# Integrative Genomics Analysis Reveals a Novel 21q22.11 Locus Contributing to Susceptibility of COVID-19

**DOI:** 10.1101/2020.09.16.20195685

**Authors:** Yunlong Ma, Yukuan Huang, Sen Zhao, Yinghao Yao, Yaru Zhang, Jia Qu, Nan Wu, Jianzhong Su

## Abstract

The systematic identification of host genetic risk factors is essential for the understanding and treatment of COVID-19. By performing a meta-analysis of two independent genome-wide association (GWAS) summary datasets (N = 680,128), a novel locus at 21q22.11 was identified to be associated with COVID-19 infection (rs9976829 in *IFNAR2* and upstream of *IL10RB*, OR = 1.16, 95% CI = 1.09 - 1.23, P = 2.57×10^−6^). The rs9976829 represents a strong splicing quantitative trait locus (sQTL) for both *IFNAR2* and *IL10RB* genes, especially in lung tissue (P = 1.8×10^−24^). Gene-based association analysis also found *IFNAR2* was significantly associated with COVID-19 infection (P = 2.58×10^−7^). Integrative genomics analysis of combining GWAS with eQTL data showed the expression variations of *IFNAR2* and *IL10RB* have prominent effects on COVID-19 in various types of tissues, especially in lung tissue. The majority of *IFNAR2*-expressing cells were dendritic cells (40%) and plasmacytoid dendritic cells (38.5%), and *IL10RB*-expressing cells were mainly nonclassical monocytes (29.6%). *IFNAR2* and *IL10RB* are targeted by several interferons-related drugs. Together, our results uncover 21q22.11 as a novel susceptibility locus for COVID-19, in which individuals with G alleles of rs9976829 have a higher probability of COVID-19 susceptibility than those with non-G alleles.

## 1. Introduction

Coronavirus disease 2019 (COVID-19) has rapidly evolved into a global pandemic ^[1]^. The health and economy systems of most nations worldwide are suffering from severe disruptions ^[2]^. As of July 13^th^, 2020, there were more than 12.9 million confirmed patients worldwide with more than 550,000 deaths ^[3]^. The clinical manifestations of COVID-19 range from asymptomatic to severe respiratory failure ^[4]^. Early studies on COVID-19 infection have concentrated on epidemiology ^[5]^, clinical characteristics ^[6]^, and genomic features of virus ^[7]^. Understanding host genetic factors contributing to COVID-19 susceptibility is essential for the precise management in the community.

Recently, a growing number of researchers have concentrated on the involvement of host genetic factors in COVID-19. Through performing a genome-wide association study (GWAS) with 1,610 severe COVID-19 patients and 2,205 controls, Ellinghaus et al. ^[8]^ reported two important gene clusters of 3p21.31 and 9q34.2 as genetic susceptibility loci for severe COVID-19, and confirmed a potential involvement of the ABO blood-group system. From a population perspective, the COVID-19 Host Genetic Consortium launched the “COVID-19 Host Genetics Initiative” to collect data from the genetics community to uncover the genetic determinants of COVID-19 susceptibility, severity, and outcomes ^[2]^. However, identification of more host genetic risk factors is limited by the sample size of a single study.

Here, we performed a meta-analysis by combining two independent GWAS summary statistics with a large-scale sample size to identify novel genes for COVID-19 susceptibility. The systematic bioinformatics analyses were performed, including MAGMA gene-based association analysis, S-PrediXcan and S-MultiXcan analysis, Sherlock-based inference analysis, gene-property analysis, single-cell RNA analysis, and functional enrichment analysis. We uncover the genes and pathways convey risk of COVID-19 infection and give a clue of the potential effective drugs for treating COVID-19.

## 2. Results

### 2.1. SNP-level association analysis reveals a novel susceptible locus 21q22.11 for COVID-19

By conducting a meta-analysis of GWAS summary data from Ellinghaus et al. ^[8]^ (COVID_I: 1,610 COVID-19 patients and 2,205 controls) and the COVID-19 Host Genetic Consortium ^[2]^ (ANA5: 1,678 COVID-19 patients and 674,635 controls), we confirmed two reported loci of 3p21.31 and 9q34.2 to be associated with COVID-19 infection (rs11385942 in *SLC6A20*, P = 2.87×10^−16^, and rs8176719 in *ABO*, P = 4×10^−7^; Figure 1, Table 1, and Supplemental Figures S1-S2). The reported rs657152 in *ABO*, which is high linkage disequilibrium with rs8176719, remains suggestively significant (P = 5.53×10^−6^). Notably, we identified a novel locus at 21q22.11 to be associated with COVID-19 infection (rs9976829 in *IFNAR2-IL10RB*, OR = 1.16, 95% CI = 1.09 - 1.23, P =2.57×10^−6^; Table 1 and Figure 1). The rs9976829 represents a splicing quantitative trait locus (sQTL) for both *IFNAR2* and *IL10RB* genes across multiple tissues with the strongest significance in the lung tissue (P = 1.8×10^−24^; Supplemental Figure S3).

**Table 1.**
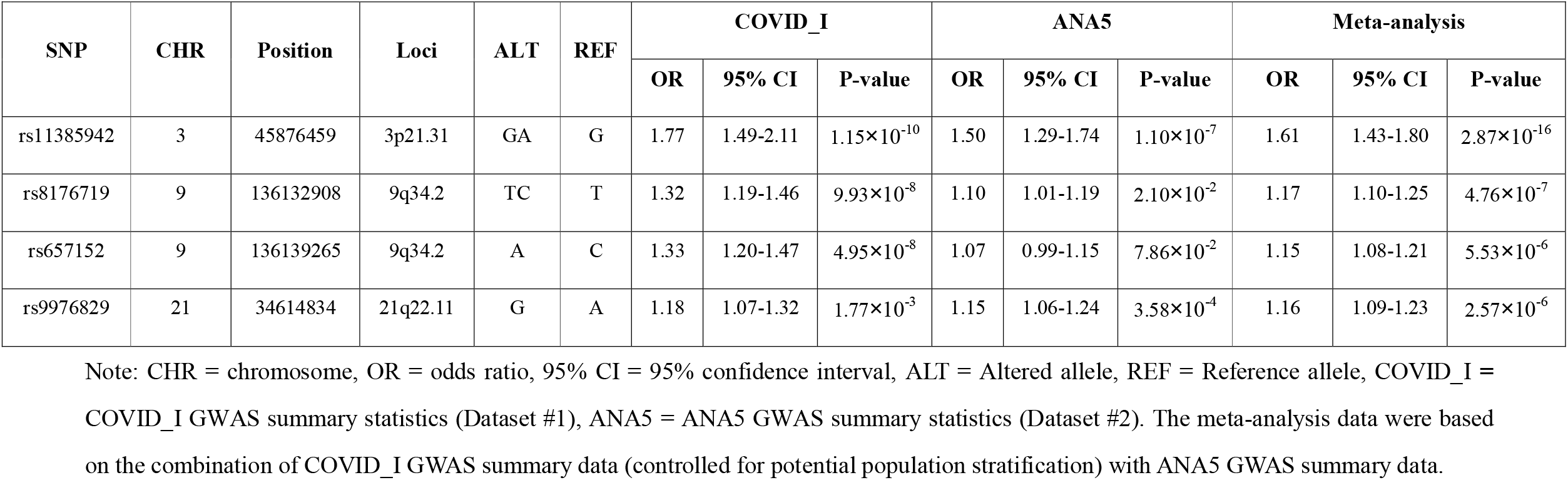
Susceptibility loci associated with COVID-19 identified by meta-analysis of GWAS summary data.

| SNP | CHR | Position | Loci | ALT | REF | COVID_I |  |  | ANA5 |  |  | Meta-analysis |  |  |
| --- | --- | --- | --- | --- | --- | --- | --- | --- | --- | --- | --- | --- | --- | --- |
|  |  |  |  |  |  | OR | 95% CI | P-value | OR | 95% CI | P-value | OR | 95% CI | P-value |
| rs11385942 | 3 | 45876459 | 3p21.31 | GA | G | 1.77 | 1.49-2.11 | $1.15 \times 10^{-10}$ | 1.50 | 1.29-1.74 | $1.10 \times 10^{-7}$ | 1.61 | 1.43-1.80 | $2.87 \times 10^{-16}$ |
| rs8176719 | 9 | 136132908 | 9q34.2 | TC | T | 1.32 | 1.19-1.46 | $9.93 \times 10^{-8}$ | 1.10 | 1.01-1.19 | $2.10 \times 10^{-2}$ | 1.17 | 1.10-1.25 | $4.76 \times 10^{-7}$ |
| rs657152 | 9 | 136139265 | 9q34.2 | A | C | 1.33 | 1.20-1.47 | $4.95 \times 10^{-8}$ | 1.07 | 0.99-1.15 | $7.86 \times 10^{-2}$ | 1.15 | 1.08-1.21 | $5.53 \times 10^{-6}$ |
| rs9976829 | 21 | 34614834 | 21q22.11 | G | A | 1.18 | 1.07-1.32 | $1.77 \times 10^{-3}$ | 1.15 | 1.06-1.24 | $3.58 \times 10^{-4}$ | 1.16 | 1.09-1.23 | $2.57 \times 10^{-6}$ |

Note: CHR = chromosome, OR = odds ratio, 95% CI = 95% confidence interval, ALT = Altered allele, REF = Reference allele, COVID\_I = COVID\_I GWAS summary statistics (Dataset #1), ANA5 = ANA5 GWAS summary statistics (Dataset #2). The meta-analysis data were based on the combination of COVID\_I GWAS summary data (controlled for potential population stratification) with ANA5 GWAS summary data.

**Figure 1.**
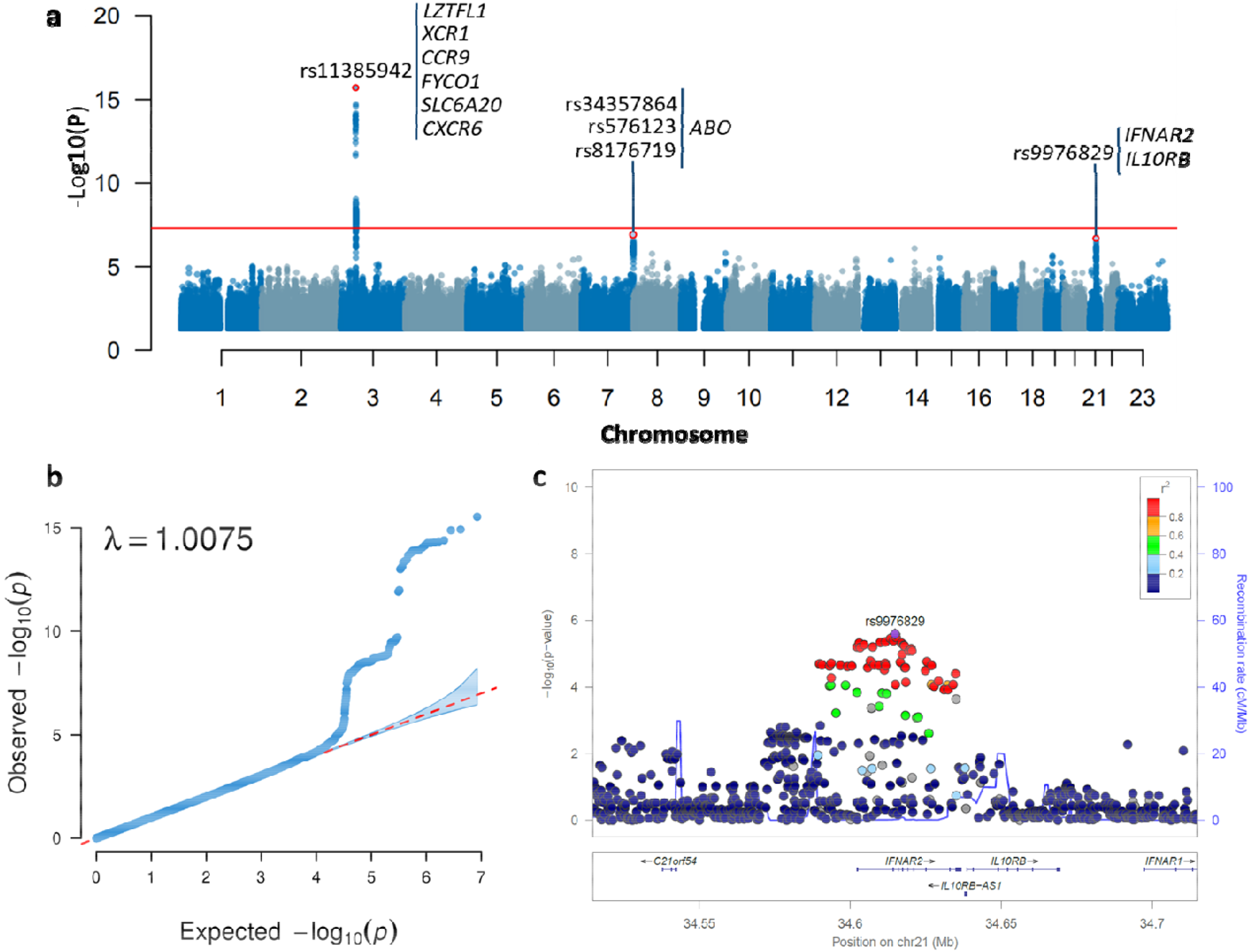
Meta-analysis of GWAS summary data highlighting susceptibility loci for COVID-19. a) Manhattan plot of the meta-analysis GWAS summary statistics highlighting three susceptibility loci for COVID-19. The Manhattan plot is shown of the meta-GWAS summary statistics of meta-analyzing the COVID_I GWAS data (controlled for potential population stratification) with ANA5 GWAS data. The red horizontal line marks the genome-wide significance threshold of a P value less than 5 × 10^−8^. b) Quantile-quantile (QQ) plot of the meta-analysis GWAS summary statistics. All 8,424,883 high-quality SNPs with a MAF ≥ 1% and imputation R^2^ ≥ 0.6 were used for plotting. In QQ plot the 2.5^th^ and 97.5^th^ centiles of the distribution under random sampling and the null hypothesis form the 95% concentration band. The genomic inflation factor lambda (λ) is 1.0075. c) Regional association plot for 21q22.11 locus of meta-GWAS summary statistics. Regional association plot is shown for 21q22.11 locus of the meta-GWAS summary statistics of meta-analyzing the COVID_I GWAS data (controlled for potential population stratification) with ANA5 GWAS data. The purple diamond marks the most strongly associated SNP of rs9976829 with COVID-19. The color illustrates LD information with rs9976829, as shown in the color legend.

### 2.2. Gene-based association analysis identifies nine risk genes for COVID-19

We performed a MAGMA gene-based association analysis by using the meta-GWAS results and found that nine genes in three loci of 3p21.31 (*LZTFL1, XCR1, CCR9, FYCO1, SLC6A20*, and *CXCR6*), 9q21.32 (*HNRNPK* and *RMI1*), and 21q22.11 (*IFNAR2*) were significantly associated with COVID-19 infection (FDR < 0.05; Figure 2 and Table 2). As expected, the gene of *ABO* showed a nominally significant association with COVID-19 (P = 6.55×10^−4^) consistent with the previous report ^[8]^. Of note, *IFNAR2* (P = 2.58×10^−7^), *HNRNPK* (P = 1.46×10^−5^), and *RMI1* (P = 1.86×10^−5^) were identified for the first time to be associated with COVID-19 infection. There were other 30 genes showing suggestive associations with COVID-19 (P < 1 × 10^−3^, Supplemental Table S1). Meanwhile, we performed two MAGMA gene-based association analyses for COVID_I GWAS summary data from Ellinghaus et al. ^[8]^ and the COVID-19 Host Genetic Consortium (ANA5) ^[2]^, respectively. We found that these nine identified genes showed significant or suggestive associations with COVID-19 in both COVID-I and ANA5 datasets (Table 2), indicating that our meta-analysis based on larger samples enhance the statistical power to uncover risk genes for COVID-19.

**Table 2.**
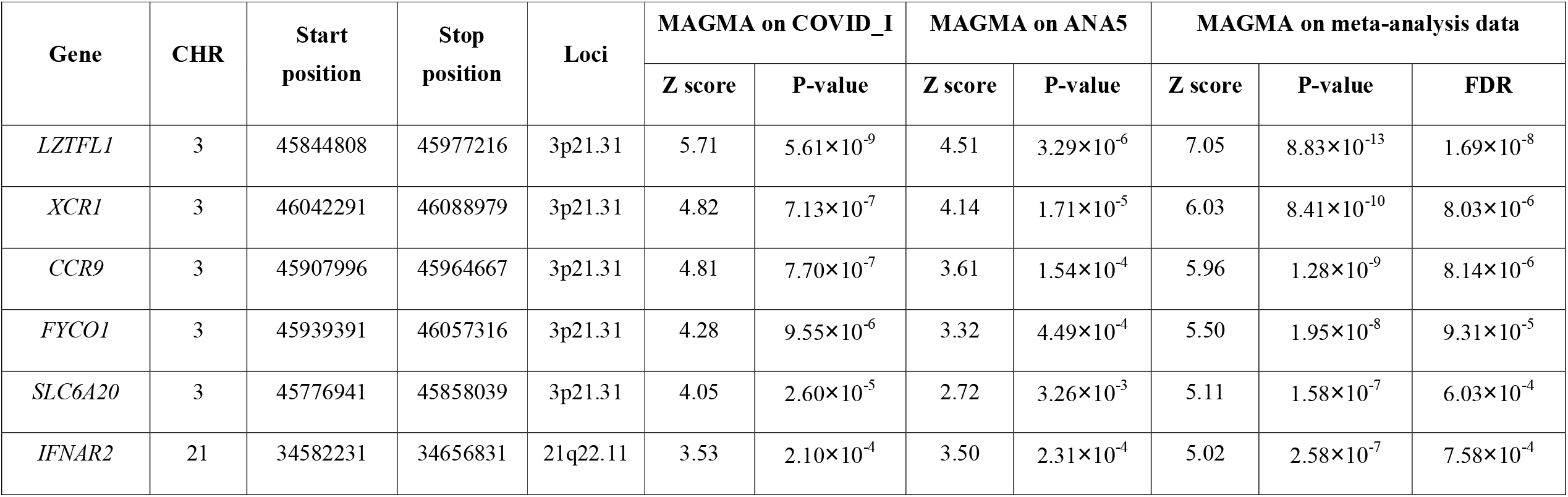
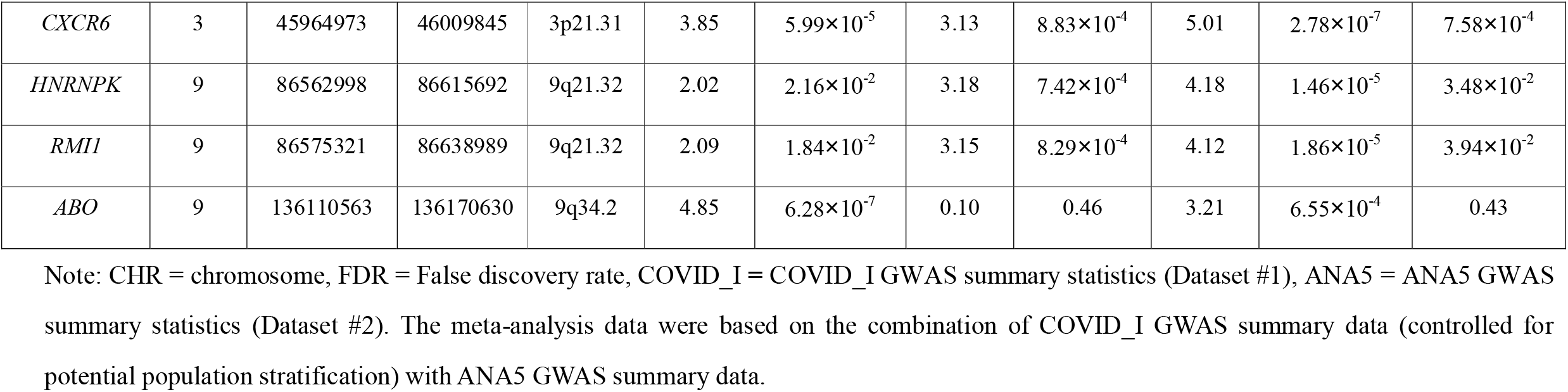
Significant genes associated with Covid-19 identified by MAGMA gene-based association analysis.

**Figure 2.**
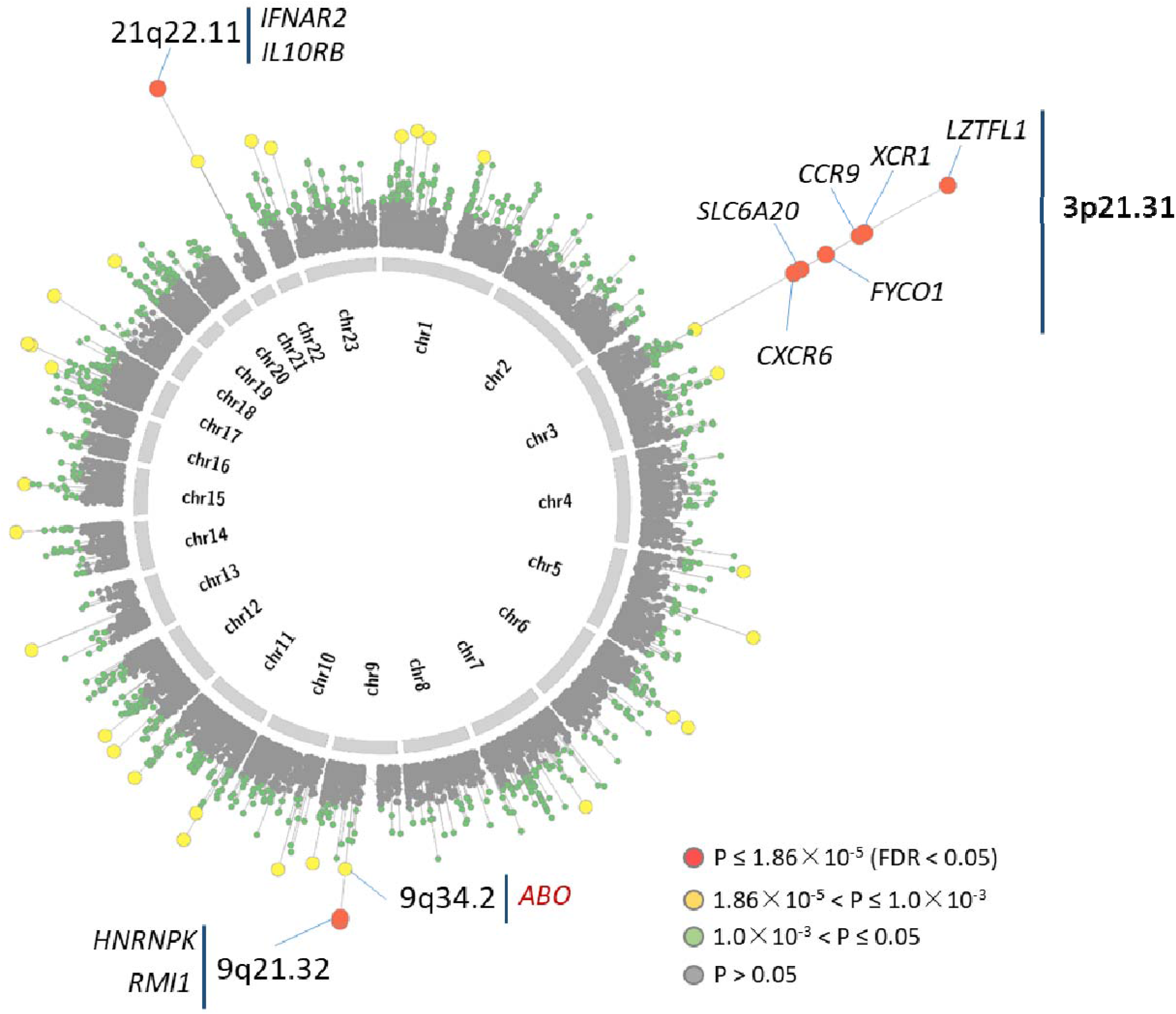
Circus plot showed the results of gene-based association analysis. Note: The inner ring shows the 22 autosomal human chromosomes (Chr1-22) and X chromosome (Chr23). A circular symbol in the outer ring represents a gene. Color indicates the statistical significance of genes (red marks genes significantly associated with COVID-19 with FDR < 0.05, yellow indicates genes with 1.86×10^−5^ < P ≤ 1×10^−3^, green marks genes with 1×10^−3^ < P ≤ 0.05, and gray represents genes with P > 0.05).

### 2.3. Cytokine-related pathways enriched by risk genes for COVID-19

As mentioned above, there were 41 genes showing significant or suggestive associations with COVID-19 (P < 1 × 10^−3^, Supplemental Table S1). We performed a pathway enrichment analysis of these 41 identified genes for COVID-19 susceptibility (Methods), and found they were significantly enriched in two pathways of cytokine-cytokine receptor interaction (FDR = 0.009) and chemokine signaling pathway (FDR = 0.009) (Figure 3a). Additionally, there were seven pathways showing suggestive associations (P < 0.05; Supplemental Table S2), including Kaposi sarcoma-associated herpesvirus infection (P = 0.0086), human cytomegalovirus infection (P = 0.014), and human papilloma virus infection (P = 0.027). Meanwhile, we conducted a GO enrichment analysis and found 3 GO-terms were significantly overrepresented (FDR < 0.05; Figure 3b and Supplemental Table S3). That is, cytokine receptor activity (FDR = 1.01 × 10^−5^), cytokine binding (FDR = 0.022), and peptide receptor activity (FDR = 0.027). Previous studies demonstrated that soluble cytokines activate an anti-viral and anti-proliferate state by inducing the expression of many interferon-stimulated genes to prevent viral replication ^[9]^. Network-based enrichment analysis showed that these 41 genes are at least partially biologically connected (P = 0.013, Supplemental Figure S8). These results suggest cytokine-related pathways or functional terms may play important roles in the process of COVID-19 infection.

**Figure 3.**
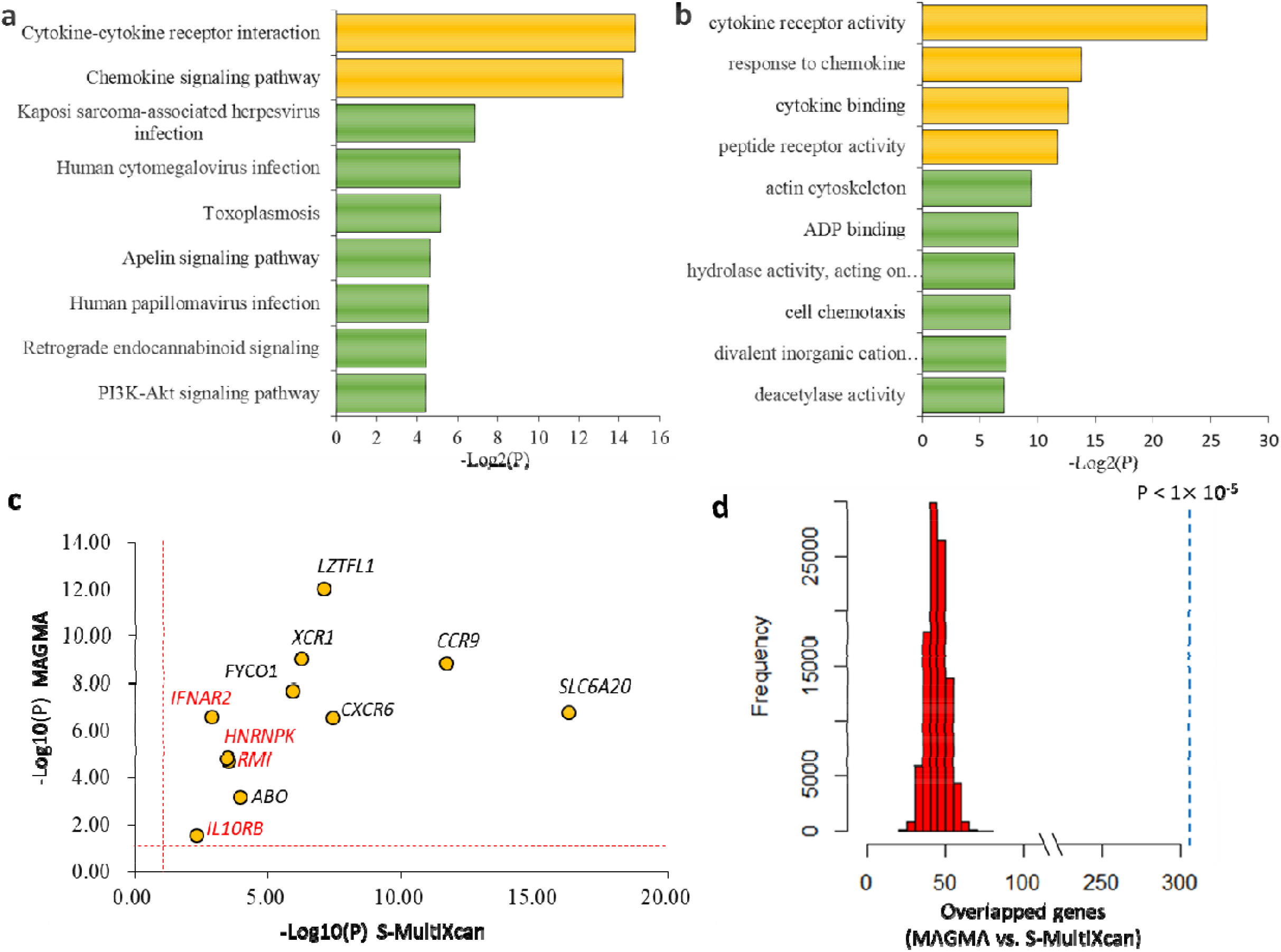
Functional enrichment analysis of genes associated with COVID-19. a) Pathway enrichment analysis identified 9 significant or suggestive KEGG pathways enriched by COVID-19-associated genes. b) GO enrichment analysis identified 10 significant or suggestive GO-terms enriched by COVID-19-associated genes. a) - b) The green bar represents a suggestive enrichment (P < 0.05), and the orange bar represents a significant enrichment (FDR < 0.05). c) Scatter plot show the consistency of 11 risk genes identified from both MAGMA and S-MultiXcan analysis. The vertical and horizontal dotted lines represent -log10 (P = 0.05). d) *In silico* permutation analysis of 100,000 times of random selections. This permutation analysis was used to compare the overlapped genes between MAGMA and S-MultiXcan (see Methods). The empirical P value is less than 1×10^−5^.

### 2.4. Expression of *IFNAR2* and *IL10RB* associated with COVID-19 across multiple tissues

To highlight the functional association of these 11 identified genes with COVID-19, we conducted two independent integrative genomics analyses by incorporating meta-GWAS summary data with eQTL data across 49 GTEx tissues. Using S-PrediXcan analysis, we found that the expression variations of these 11 genes have prominent effects on COVID-19 in various types of tissues (Supplemental Table S5). Consistently, Sherlock-based integrative genomics analysis showed seven genes including *IFNAR2* and *IL10RB* whose genetically regulated expression were significantly associated with COVID-19 across multiple tissues (Supplemental Table S6). For example, the *IFNAR2* gene was associated with COVID-19 across six tissues including lung (P = 2.44 × 10^−4^). The *IL10RB* gene showed associations with COVID-19 across 24 tissues with the most significant tissue of cells-transformed fibroblasts (P = 2.80 × 10^−5^).

We further used S-MultiXcan to meta-analyze the tissue-specific associations from S-PrediXcan across 49 GTEx tissues, and found these 11 genes whose expression were significantly associated with COVID-19, which support our results from the MAGMA analysis (Figure 3c and Supplemental Table S7). Additionally, our *in silico* permutation analysis also showed that genes identified from MAGMA analysis had a significantly higher overlap with S-MultiXcan-identified genes than random events (P < 1 × 10^−5^; Figure 3d).

### 2.5. *IFNAR2* and *IL10RB* specially expressed in immunity associated cell types in lung tissue

To examine the links between tissue-specific gene expression profiles and COVID-19 gene associations, we conducted a MAGMA gene-property analysis in 53 specific tissue types and 30 general tissue types. We found that the association signals were enriched in lung, thyroid, and esophagus tissue in 30 general tissue types (Supplemental Figure S4a). In the analysis of 53 specific tissues, COVID-19 gene associations were also enriched in lung, cultured fibroblasts, and thyroid (Supplemental Figure S4b). These results suggest that these identified genes for COVID-19 may have important functions in lung tissue.

Using 50 cell populations across four compartments (epithelial, endothelial, stromal, and immune) of lung tissue, we identified the primarily expressed cells of these 11 risk genes associated with COVID-19 (Figure 4, Supplemental Table S4, and Supplemental Figure S5). The *HNRNPK* gene was widely expressed in various cell types across all four compartments (Supplemental Figure S6). However, the majority of *IFNAR2*-expressing cells were dendritic cells (40%) and plasmacytoid dendritic cells (38.5%) (Figure 4). The *IFNAR2* was also expressed within lipofibroblast (20%), ciliated (19.1%) and nonclassical monocyte (18.5%), albeit at diminished abundance compared with dendritic cells; *IL10RB* was primarily expressed within nonclassical monocyte (29.6%). Plasmacytoid dendritic cells produce large amounts of type I interferons-proteins that are important for immunity to viruses ^[10]^. Our data indicated that *IFNAR2* and *IL10RB* could play regulatory roles in the pulmonary immune response.

**Figure 4.**
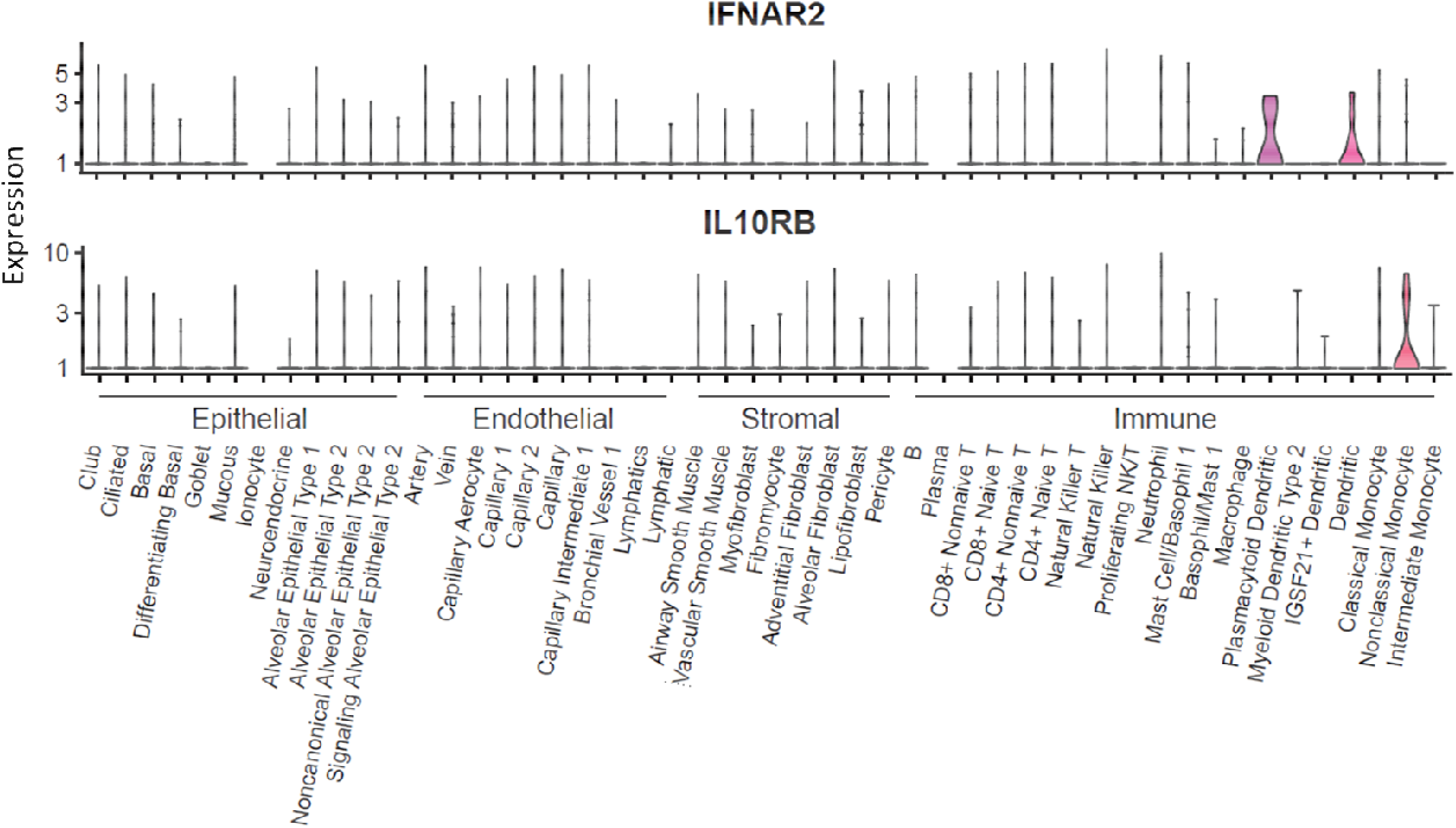
Expression of *IFNAR2* and *IL10RB* among 50 cellular populations from lung tissue. This plot is based on a data set as a part of the Human Lung Atlas, consisting of 50 cell populations across 4 compartments (epithelial, endothelial, stromal, and immune) of lung tissue (x axis). y axis represents the expression level with log transformed count.

### 2.6. Potential drugs targeted with *IFNAR2* and *IL10RB*

By performing a drug-gene interaction analysis and literature mining, we found that seven of 11 COVID-19-associated genes (63.6%) were enriched in five potential “druggable” gene categories (Supplemental Table S8 and Supplemental Figure S7). The gene of *CCR9* is targeted by two FDA-approved drugs including hydroxyurea and hydralazine (Supplemental Table S9). There are nine FDA-approved drugs showing agonist-receptor interactions with *IFNAR2* (Supplemental Table S9), including interferon alfa-2a, interferon alfacon-1, and interferon beta-1a, which could be useful alone or in combination with other antiviral drugs for treating SARS-CoV infection ^[11]^. Loutfy et al. ^[12]^ reported that interferon alfacon-1 plus corticosteroids showed association with improved oxygen saturation and more rapid resolution of radiographic lung opacities than systemic corticosteroid alone in severe acute respiratory syndrome (SARS). Treatment with interferon has shown preliminary benefits for patients with COVID-19 ^[13]^ and is being evaluated by numerous ongoing clinical trials. The IFN-beta treatment could effectively block SARS-CoV-2 replication ^[14]^. The *IL10RB* gene is targeted by peginterferon lambda-1a (Supplemental Table S9), which has been designed to treat mild COVID-19 in a clinical trial (identifier: NCT04331899). These results suggest that interferons associated with *IFNAR2* and *IL10RB* exert potential effects on the treatment of COVID-19.

## 3. Discussion

Using a meta-analytic method to combine two existing GWAS summary datasets with a large-scale sample size, we validated two reported genetic loci on chromosome 3p21.31 and 9q34.2 to be significantly associated with COVID-19 infection, and found a novel locus at a chromosome 21q22.11 gene cluster conveying susceptibility of COVID-19.

On chromosome 21q22.11, the peak association signal covered two genes of *IFNAR2* and *IL10RB*, which have biological functions that probably related to COVID-19. *IFNAR2* encodes a type I membrane protein, which forms one of the two chains of a receptor for interferons alpha and beta. The deficiency of IFNAR2 supports an essential role for interferons alpha and beta in human antiviral immunity ^[15]^. Notably, dysregulation of type I interferon response has been observed in COVID-19 patients ^[16]^. Impaired type I interferon activity in the blood could be a hallmark of severe COVID-19 ^[17]^. Furthermore, *IFNAR2* is required for anti-influenza immunity and related to the risk of post-influenza bacterial superinfections ^[18]^. As for *IL10RB*, its encoded protein belongs to the cytokine receptor family and is an accessory chain essential for the active interleukin 10 receptor complex. Variants in *IFNAR2* and *IL10RB* gene were associated with the susceptibility to hepatitis B virus (HBV) infection ^[19]^. Most recently, the interferon pathway is identified to be targeted by the COVID-19 viral protein of Nsp13 ^[20]^.

Functional enrichment analysis showed that cytokine-related pathways including cytokine receptor activity, cytokine-cytokine receptor interaction, and chemokine signaling pathway were significantly enriched by genes associated with COVID-19. Seven risk genes of *IFNAR2, IL10RB, XCR1, CXCR6, CCR9, CCR1*, and *GNG12* have implicated in these pathways. There were four genes encoding chemokine receptors, including the X-C motif chemokine receptor 1 (XCR1), the C-×-C motif chemokine receptor 6 (CXCR6), the CC motif chemokine receptor 9 (CCR9), and the CC motif chemokine receptor 1 (CCR1). Vaccine molecules targeting XCR1 on cross-presenting dendritic cells enhance a protective CD8+ T-cell responses against influenza virus ^[21]^. CXCR6 modulates the localization of lung tissue-resident memory CD8+ T-cells throughout the sustained immune response to respiratory pathogens, including influenza viruses ^[22]^. Both CCR9 and CCR1 also have related functions on immune response to respiratory influenza infection ^[23]^. Varied manifestations in COVID-19 infection may result from different host genetic factors, which are probably related to immune response ^[24]^. Through inducing the expression of many interferon-stimulated genes, soluble cytokines have anti-viral, anti-proliferate, and immunomodulatory effects on obstructing viral replication ^[9]^. Together, cytokine-related pathways potentially play important roles in the pathogenesis of COVID-19 infection, and more relevant studies should be performed to explore the underlying biological mechanisms.

With regard to the novel gene of *HNRNPK*, it belongs to the subfamily of ubiquitously expressed heterogeneous nuclear ribonucleoproteins (hnRNP), of which proteins have important roles in cell cycle progression. *HNRNPK* acts as a central hub in the replication cycle of multiple viruses including HCV ^[25]^. An interaction of *HNRNPK* and *HNRNPA2B1* with hepatitis E virus (HEV) promoters has important roles in HEV replication ^[26]^. Two cellular RNA binding proteins of hnRNPK and NS1-BP have important roles in regulating influenza A virus RNA splicing ^[27]^. As for the *RMI1* gene, its protein is an essential component of the RMI complex, which has a crucial role in DNA repair and maintaining genome stability ^[28]^. Since there existed important functions of these identified genes, molecular studies are warrant to illustrate the functional consequences of detected association signals.

The power of our study is limited by the difference in study design between the two datasets we analyzed: the COVID-19 Host Genetic Consortium included patients with mild or severe COVID-19 but the Ellinghaus et al. ^[8]^ study only included severe COVID-19. Because most of the COVID-19 infected individuals are asymptomatic, population-based controls used in the original studies may contain a substantial proportion of asymptomatic patients, which further reduced the power of our meta-analysis. Due to data from the two included GWASs were based on summary statistics, the population stratification was not assessed in the current meta-analysis. For the Ellinghaus et al. ^[8]^ study, to examine for population stratification within and across Italian and Spanish panels, a principal component analysis (PCA) was performed by using the FlashPCA ^[29]^. Covariates from 10 PCA were conducted to control for potential population stratification (COVID_I). As for the ANA5 GWAS summary data from the COVID-19 Host Genetic Consortium constructed by 10 contributing studies, the strategy of population stratification is unknown.

## 4. Conclusions

In summary, our findings uncover 21q22.11 as a novel risk locus for COVID-19 susceptibility and implicate the potential role of interferons targeting *IFNAR2* and *IL10RB* in the treatment of COVID-19. Individuals with the G alleles of rs9976829 have a 16% greater chance of COVID-19 infection compared with these carrying no such allele. The efficacy and the safety of interferon products are still being evaluated by numerous ongoing clinical trials, which may be strengthened by subgrouping the patients according to their genotypes of the *IFNAR2* and the *IL10RB* loci. Further studies are needed to delineate current findings and understand the underlying pathophysiology of COVID-19.

## 5. Methods

### GWAS summary data from Ellinghaus et al. (Dataset #1)

For this GWAS recently reported by Ellinghaus et al. ^[8]^, there were 1,980 patients with severe COVID-19 enrolled from seven hospitals in the Italian and Spanish epicenters of the SARS-CoV-2 pandemic in the Europe. A total of 2,381 control participants were enrolled from Italy and Spain. After stringent quality control and excluding population outliers, 1,610 patients with COVID-19 with respiratory failure (835 Italian and 775 Spanish COVID-19 cases) and 2,205 control participants (1,255 Italian and 950 Spanish controls) were included in the final GWAS. In total, 8,965,091 high-quality SNPs (post imputation R^2^≥ 0.6 and minor allele frequency (MAF) ≥ 1%) were included in the Italian cohort and 9,140,716 high-quality SNPs in the Spanish cohort. The GWAS summary statistics (COVID_I) are publicly available in the website (www.c19-genetics.eu). For more detailed information, please refer to the original article ^[8]^.

### GWAS summary data from the COVID-19 Host Genetic Consortium (Dataset #2)

This GWAS summary statistics of the publicly available COVID-19 HGI GWAS meta-analyses round 2 (ANA5, susceptibility [affected vs. population]) was downloaded from the official website of the COVID-19 Host Genetic Consortium ^[2]^ (www.covid19hg.org/results; analysis named “20200508-results-ANA5_ALL_inv_var_meta”; file named “COVID19_HGI_ANA5_20200513.txt.gz”; release date of May 15 2020). There were 1,678 COVID-19 patients and 674,635 control participants from 10 contributing studies. For the GWAS summary statistics, there were a total of 34,010,457 genetic variants included with a MAF threshold of 0.0001 and an imputation score filter of 0.6. For more detailed information, please refer to the original article ^[2]^.

### Meta-analysis of GWAS summary data

By using the meta-analysis tool of METAL ^[30]^, a fixed-effects meta-analysis was performed to identify risk genes for COVID-19 across two GWAS summary datasets (Dataset #1: COVID_I GWAS summary statistics, and Dataset #2: ANA5 GWAS summary statistics). After removing low-quality and non-matched SNPs, there were 8,424,883 high-quality SNPs with a MAF ≥ 1% and ≥ imputation R^2^ 0.6 that were common to both datasets with the use of effect-size estimates (BETA) and their standard errors (SE) for the meta-analysis. With regard to the genome-wide meta-analysis, we adopted the widely-used threshold of 5×10^−8^ for combined P values to determine statistical significance. As reported in Ellinghaus et al. ^[8]^, we used the combined P value and combined effect (E) with its SE generated by the METAL to compute the odds ratio (OR) and its 95% confidence interval (CI): 1) OR = exp (E); 2) the upper confidence limit (OR_95 U) = exp (E + 1.96*SE); 3) the lower confidence limit (OR_95L) = exp (E - 1.96*SE). The *qqman* package in R platform was used to generate Manhattan plot and quantile-quantile (QQ) plot. The web-access tool of *LocusZoom* ^[31]^ was used to visualize regional association plots (http://locuszoom.sph.umich.edu/).

### Gene-based association analysis

We conducted a gene-based association analysis of our meta-GWAS summary data for COVID-19 by using the Multi-marker Analysis of GenoMic Annotation (MAGMA) ^[32]^, which utilizes a multiple regression method to identify multi-marker aggregated effects that account for SNP P values and linkage disequilibrium (LD) between SNPs. The analyzed SNP set of each gene was based on whether the SNP located in the gene body region or within extended +/- 20 kb downstream or upstream of the gene. The LDs among SNPs were calculated based on the 1,000 Genomes Phases 3 European Panel ^[33]^. The Benjamini-Hochberg false discovery rate (FDR) method was used to correct the association results for multiple testing. The P value threshold of 1.86 ×10^−5^ was applied.

### Functional enrichment analysis

To annotate the molecular functions and biological pathways of these COVID-19-associated genes (Supplemental Table S2), we performed a functional enrichment analysis by using the WebGestalt tool ^[34]^ based on the Kyoto Encyclopedia of Genes and Genomes (KEGG) pathways and gene ontology (GO) terms. Using the overrepresentation analysis, the WebGestalt could identify functional association between COVID-19-associated genes and KEGG pathways. Furthermore, GO enrichment analysis was performed by using 3 categories of GO terms: biological process, cellular component, and molecular function. The redundancies of GO-terms were removed. The hypergenometric test was applied to assess the statistical significance. P values were corrected for multiple testing using the FDR, and a P value threshold of 2.9 ×10^−4^ was applied.

### Gene-property analysis

We performed a MAGMA gene-property analysis ^[32]^, which is implemented in FUMA ^[35]^. The gene expression data from 83 tissues GTEx RNA-seq data (version 8) were used to parse the gene expression profiles. Expression values (TPM) were log2 transformed and average expression values were adopted per tissue. The gene-property analysis was conducted for 53 specific tissue types and 30 general tissue types, respectively. Bonferroni correction for multiple testing was used for the examined tissue types.

### Single-cell RNA-seq analysis for lung tissue

We performed single-cell data analysis from normal lung tissue sequenced by using Smart-Seq2. This data set is a part of the Human Lung Atlas ^[36]^, consisting of 50 cell populations across 4 compartments (epithelial, endothelial, stromal, and immune) of lung tissue. All 9,404 cells with distinct cellular identities were download from the Human Lung Atlas. Cells from 3 donors with FACS-sorted strategy were available at the Synapse (accession numbers: syn22168639, syn22168625, and syn22168622).

### S-PrediXcan and S-MultiXcan analysis

We applied S-PrediXcan ^[37]^ to integrate expression quantitative trait loci (eQTL) data with genetic associations from GWAS summary statistics to identify genes, which genetically predicted expression levels are associated with COVID-19. S-PrediXcan firstly estimates gene expression weights by training a linear prediction model (MASHR model) in samples with both SNP genotype and gene expression data. These estimated weights are processed with beta values and standard errors from meta-GWAS summary data on COVID-19 to predict gene expression from GWAS summary statistics, while combining the variance and co-variance of SNPs from an LD reference panel based on the 1000 Genomes Project Phase 3 genotypes ^[33]^. The eQTL data for 49 tissues from the GTEx Project (version 8) were used in the current analysis. S-PrediXcan was performed for each of 49 tissues for a total of 659,158 gene-tissue pairs. To increase power to identify genes whose expression is similarly differentially regulated across tissues, we meta-analyzed the S-PrediXcan results with the use of the S-MultiXcan method ^[38]^, which employs multivariate regression to integrate evidence across 49 GTEx tissues with a total of 22,327 genes. Significant associations were determined by using Bonferroni correction.

### Sherlock-based integrative genomics analysis

To further validate these identified host genes for COVID-19, we also applied an independent approach of Sherlock-based integrative genomics analysis based on a Bayesian inference algorithm ^[39]^. The Sherlock-based integrative analysis incorporates genetic information from meta-GWAS summary statistics on COVID-19 with eQTL data across 49 tissues from the GTEx Project (version 7) to prioritize important risk genes. It should be noted that Sherlock employs different algorithm and strategies to perform the statistical inference compared with S-PrediXcan. Briefly, Sherlock first searches expression-associated SNPs (named as eSNPs) across different GTEx tissues. Then, Sherlock estimates the possible association of eSNPs with COVID-19 using our meta-GWAS summary data. Sherlock computes individual Logarithm of the Bayes Factor (LBF) for each SNP pair, and the sum of these constitutes the final LBF score for each gene. There are three potential scenarios: 1) if an eSNP in a given gene showed a significant association with COVID-19, a positive score would be assigned; 2) if an eSNP in a given gene showed a non-significant association with COVID-19, a negative score would be assigned; 3) no score was assigned if an SNP was not eSNP but showed a significant association with COVID-19. The Sherlock applied the simulation analysis to compute the p value of the Bayes factor for each gene, as reference of a method of Bayes/non-Bayes compromise ^[40]^. We used the Bonferroni correction method to correct the significance for multiple testing. Due to we tested the Sherlock analysis in 49 GTEx tissues, there existed different thresholds across tissues. For example, the P value threshold was 6.64 ×10^−6^ (0.05/7,529) for lung tissue.

### In silico permutation analysis

To determine whether there exist a higher overlapped rate of genes identified from MAGMA analysis (Gene set #1: N = 1,005, P < 0.05) with genes from S-MultiXcan analysis (Gene set #2: N = 1,141, P < 0.05) than that from random selections, we conducted a computer-based permutation analysis of 100,000 times of random selections ^[41]^. First, we counted the overlapped genes between Gen sets #1 and #2 (*N* _observation_). Secondly, we used all tested genes from S-MultiXcan analysis as background genes (*N* _background_ = 22,326 genes). Then, through randomly selecting the same number of genes as Gene set #2 from background genes to compare with Gene set #1 for 100,000 times, we counted the number of overlapped genes in each time (*N* _random_). In the third step, we summed the counts of *N* _observation_ ≤ *N* _random_ and divided by 100,000 to calculate empirically permuted P value. P < 0.05 is considered to be significant.

### Drug-gene interaction analysis

We submitted these 11 COVID-19-associated genes into the widely-used Drug Gene Interaction Database (DGIdb v.3.0.2; http://www.dgidb.org/) to identify drug-gene interactions with Food and Drug Administration (FDA)-approved pharmaceutical compounds as well as antineoplastic and immunotherapies drugs depended on 20 databases with 51 known interaction types, and search 10 databases to find genes with potential drug abilities.

## Data Availability

All the GWAS summary statistics used in this study can be accessed in the official websites (www.covid19hg.org/results and www.c19-genetics.eu). The GTEx eQTL data (version 8) were downloaded from Zenodo repository (https://zenodo.org/record/3518299#.Xv6Z6igzbgl). All analysis code in the Methods is available in a publicly available GitHub repository at https://github.com/YukuanHuang/Host_genetics_for_COVID-19.

https://www.covid19hg.org

https://www.c19-genetics.eu

https://zenodo.org/record/3518299#.Xv6Z6igzbgl

## Supporting Information

Supporting Information is available from the Wiley Online Library or from the author.

## Acknowledgements

We thanks to the helpful suggestions of Dr. Guijun Zhang. We appreciate all the authors from the COVID-19 Host Genetic Consortium, as well as Ellinghaus and his colleagues who have deposited and shared GWAS summary data on public databases. This study was funded by the National Natural Science Foundation of China (61871294 to J.S., 81501852 to N.W.), and Science Foundation of Zhejiang Province (LR19C060001 to J.S). All analysis codes in the Methods are available in a publicly available GitHub repository at https://github.com/YukuanHuang/Host_genetics_for_COVID-19.

## Authors’ contributions

J.S., N.W., Y.M. and J.Q. conceived and designed the study. Y.M., Y.H., S.Z., G.Q. and J.Z. contributed to management of data collection. Y.M., Y.H., G.Q., J.Z., J.Q., Z.M., J.Y., J.S., N.W. and S.Z. conducted bioinformatics analysis and data interpretation. Y.M., J.S., N.W. and S.Z. wrote the manuscripts. All authors reviewed and approved the final manuscript.

## Conflict of Interest

The authors declare no conflict of interest.

## Notes

### Competing Interest Statement

The authors have declared no competing interest.

### Author Declarations

GWAS summary data of all participants used in the current study were downloaded from the public databases, which have been approved by the relevant committees in the original articles

